# MULT: An allometric body mass index (ABMI) reference to assess nutritional status of multi-ethnic children and adolescents

**DOI:** 10.1101/2024.06.07.24308574

**Authors:** Mariane Helen de Oliveira, Camila Medeiros da Silva Mazzeti, Joana Araújo, Milton Severo, Elisabete Ramos, Kévin Allan Sales Rodrigues, Débora Borges dos Santos Pereira, Wolney Lisboa Conde

**Author notes:** **Corresponding author:** / [MHO].

## Abstract

**Objectives:** To develop an allometric body mass index (ABMI) reference that adjusts the weight in relation to height, taking into account the changes during development (MULT ABMI reference), and to compare it with international BMI references.

**Methods:** The MULT ABMI reference was constructed through the LMS method, calculated with 65 644 ABMI observations of 17 447 subjects aged 5-22 years, from the United Kingdom, Ethiopia, India, Peru, Vietnam, Portugal, and Brazil. The M, S, and L curves of the MULT ABMI reference were compared with the curves of the MULT, World Health Organization (WHO), Centers for Disease Control and Prevention (CDC), International Obesity Task Force (IOTF), and Dutch Growth Study (DUTCH).

**Results:** The greater differences in the M curve between MULT ABMI and WHO, CDC, IOTF, DUTCH, and MULT BMI references were around puberty (138 to 150 months for boys; 114 to 132 for girls). MULT ABMI presented S values similar to IOTF and DUTCH BMI references for boys 60 to 114 months and then became higher, approaching the MULT BMI S values from 198 to 240 months. For girls the MULT ABMI S values were close to the IOTF, CDC, and DUTCH from 60 to 110 months, and then became higher, approaching the MULT BMI S values until 240 months.

**Conclusion:** MULT ABMI presented an advantage in comparison to the existing BMI references because it takes into account the growth changes during puberty and is a new option to assess the nutritional status of multi-ethnic populations.

## Introduction

In the last decades, the prevalence of obesity has increased among all age groups and the Body Mass Index (BMI) has been widely used as the main tool to diagnose it at a population level [1–3]. BMI is an index, calculated by dividing the body mass (BM) in kilos (kg) by the squared height in meters (m) [4,5]. This index was developed by Lambert Adolphe Jacques Quetelet in 1832 based on the assumptions that weight increases proportionally to the square of height and the fat mass (FM) and fat-free mass (FFM) scale in a similar way to weight [4–6].

Nowadays, BMI-for-age growth charts are used internationally to assess the nutritional status of children and adolescents, despite some concerns about them [7]. One issue is the that BMI is a one-dimensional index, which applies a single exponent (2) to adjust the relation between height and weight [4,8,9]. This approach may not be adequate due to the non-uniform increase in body mass and height occurred in the physical growth process and sexual maturation [10–15].

During the pubertal stage, BMI can mislead the interpretation between adiposity gain and growth spurt, indicating a need for a multidimensional approach [11,12,16]. Even Quetelet’s pioneering studies in human growth concluded that “weight gain equals the square of height”, except for growth spurts which occur after birth and during puberty [5]. It is evident that during growth, BM does not strictly adhere to the squared height relationship, which can complicate its interpretation in individuals undergoing growth [16].

In this way, research on allometric scaling has gained importance in the nutritional status assessment of children and adolescents [9,16–19]. Allometry refers to changes in the relative dimensions of body parts that correlate with changes in overall size [9]. Additionally, analysis of American population data has suggested that the relationship between BM and height can be linearized by using the quadratic transformation of height [19]. Researchers have noticed that many scale relationships could be linearized in a log-log scale through the linear equation: log y = α log x + log b [20]. Furthermore, in human allometry, there are situations where two measured variables are correlated without one directly determining the other, blurring the distinction between dependent and independent variables [21]. This is particularly evident in the relationship between BM and height, where each factor may influence the other to some degree, however the true height exponent may exceed the value of *p* determined by regression analysis [21].

The allometric scaling goal is to adjust the relationship between weight and height to represent body mass or body fat independent of height, which is the fundamental basis of the allometric body mass index (ABMI) [8,19,21]. ABMI consists in dividing the body mass (kg) by the height (m) powered by the coefficients of the allometric body mass per age (**p_t_)** [8].

At the moment, there are coefficients of allometric body mass proposed for international use [9]. These coefficients are age- and sex-specific and were calculated based on data from five countries (Brazil, United States, Mexico, South Korea, and England) [9]. However, there are no international ABMI growth charts proposed for clinical practice, even though the ABMI seems to be a formula to adjust height in relation to weight, following human growth patterns across the ages [7,9].

Therefore, this study aimed to develop an ABMI-for-age growth reference and its respective percentiles based on recent longitudinal data from multi-ethnic populations (MULT ABMI Reference) and compare it to international BMI references/percentiles of the Dutch Growth Study (DUTCH/2011), the Centers for Disease Control and Prevention (CDC/2000), the World Health Organization (WHO/2007), the International Obesity Task Force (IOTF/2012) and the MULT (2023), which is a recent developed BMI-for-age growth chart derived from multi-ethnic populations [22–26].

## Methods

### Study design and population

Young Lives (YL), Millennium Cohort Study (MCS), Adolescent Nutritional Assessment Longitudinal Study (ELANA), and Epidemiological Health Investigation of Teenagers in Porto (EPITeen) datasets were used in this study, providing longitudinal data of children and adolescents born between 1990 and 2002 [27–30].

The YL is an international study that started in 2002 with children born in Ethiopia, India (Andhra Pradesh), Peru, and Vietnam [27]. Its surveys provide demographic, socio-economic, health, cognitive, physical development, and educational data [27]. YL is composed of two cohorts: a younger cohort (YLYC) with around 8 000 children born in 2001-2002 and an older cohort (YLOC) with about 4 000 children who were born in 1994-1995 [27]. Children from the YLYC were assessed at 1, 5, 8, 12, and 15 years of age, while the ones from the YLOC were assessed at 8, 12, 15, 19, and 22 years of age [27].

The MCS provides demographic and health-related data, measures concerning child development, cognitive ability, and educational attainment of the 18 827 children born in England, Scotland, Wales, and Northern Ireland [28]. The subjects of the MCS were evaluated at 9 months and at 3, 5, 7, 11, 14, and 17 years of age, because there was no height data in the baseline, in our study, we only included their data from the second sweep forward (n = 15 588) [28].

ELANA is a study composed of 1 848 adolescents born between the 1990s and 2000s and who lived in Rio de Janeiro (Brazil) between 2010 and 2013 [29]. The State University of Rio de Janeiro (UERJ) and the Federal University of Rio de Janeiro (UFRJ) coordinated this study and its participants were from four private and two public schools in the metropolitan region of Rio de Janeiro [29]. In ELANA, there were two cohorts, one composed of middle school students (MS), and another of high school students (HS) [29]. The participants were assessed every school year, completing four evaluations for the MS and three evaluations for the HS [29].

The EPITeen is a population-based cohort of 2 942 adolescents who were born in 1990 and who attended public and private schools in the city of Porto in Portugal in 2003/2004 [30]. In addition to the recruitment at 13 years, follow-up evaluations took place at 17, 21, 24, and 27 years of age, on average [30]. During its data collection, participants’ anthropometric data at birth and during their childhood were retrieved from their health books, while additional body measure examinations were performed at all follow-ups [30,31].

Regarding the anthropometric examination, in all studies, children under two years old were measured in the supine position (length measurement), while children from two years old upward were measured in an upright position (height measurement) [27–31]. In order to assure the accuracy of the body measurement examination in the surveys, they were taken by health professionals who adhered to a standardized examination protocol [27–31].

These longitudinal studies provided data from about 32 162 participants and were conducted according to the guidelines laid down in the Declaration of Helsinki [27–30]. All participants and/or their parents or legal guardians provided written informed consent [27–30]. Data from ELANA were gathered with permission of the UFRJ and the Institute of Social Medicine’s Ethics Committee approved its study protocol (CAEE – 0020.0.259.000-09) [29,32]. Similarly, The Institute of Public Health of the University of Porto (ISPUP) authorized the retrieval of data from EPITeen, and the Ethics Committees of the Hospital São João approved the its study protocol [30,31,33].

Additionally, data from MCS and YL were gathered using the UK Data Service online platform and these studies have been approved by Ethics Committees [27,28,34–43]. The YL study protocols have subsequently received ethical committee approval before each pilot and round of data collection from both the University of Oxford and the respective ethics committees in each study country [27]. The approvals are as follows: the Central University Research Ethics Committee (CUREC) of the Social Science Division at the University of Oxford (since 2005); the Instituto de Investigación Nutricional (IIN) in Peru (since 2002); the Hanoi School of Public Health Research in Vietnam (since 2015); the Centre for Economic and Social Studies (CESS) in Hyderabad, India (since 2015); and the College of Health at Addis Ababa University in Ethiopia (since 2015) [27].

Likewise, the MCS received ethical approval for each wave of data collection from different Medical Research Ethics Committees (MREC) across the UK [28]. For MCS1, approval was granted by the South West MREC (MREC/01/6/19) in 2000/1 [28]. MCS2 was approved by the London MREC (MREC/03/2/022) in 2003/4 [28]. The MCS3 wave received approval from the London MREC Committee (05/MRE02/46) in 2005/6 [28]. For MCS4, the Yorkshire MREC provided approval (07/MRE03/32) in 2007/8 [28]. MCS5 was approved by the Yorkshire and The Humber – Leeds East MREC (11/YH/0203) in 2011/12 [28]. Finally, MCS6 received approval from the London – Central MREC (13/LO/1786) in 2015/16 [28].

### Data processing and analysis

In total, we excluded 14 715 subjects. In the selection data, firstly we excluded participants (n=177) who did not meet our eligibility requirements, which included being classified as belonging to one of the five ethnic categories (White, Black, Asian Indian, Native Peruvians, and East and Southeast Asians) and having received an assessment in all three sweeps of the ELANA HS or at least four sweeps for the other surveys.

Secondly, the following exclusions were performed: missing anthropometric and/or demographic data (n = 13 299 subjects), measurement errors, in which subjects decreased in height (≥0.5cm) over the years (n=799 subjects), and implausible values considered as height-for-age z-score below –6 standard deviations (SD) or above +6 SD or BMI-for-age z-score below –5 SD or above +5 SD (n=250 subjects) [44].

Thirdly, ABMI values with less than 10 observations per age in months (n=334) and outlier weight measurements at the population and individual levels were removed. At the population level, weight-for-age z-scores below –2 SD or above +2 SD (n= 5 672 observations) were excluded from our data pool. This outlier exclusion was performed based on WHO weight reference values for children under the age of five and according to our sample weight values for participants from five years old [45].

At the individual level, an expected BMI was calculated for each participant’s BMI data using a linear mixed-effects model [46]. This model was conducted using the *nlme* package in R, which permits layered random effects in fitting a linear mixed-effects model [46,47]. For the entire sample, BMI values that deviated below −2SD or above +2SD from the expected BMI values were excluded (n=4 727 observations). These exclusions were based on BMI values that were considered outliers within their respective subjects.

For this new growth reference for children aged 5 years and older the values of the ABMI were calculated using the formula proposed by Benn (1971), and the p_t_ coefficients, which are the exponents per month applied in the formula, were derived from the dataset compiled by Mazzeti (2018) [8,48]. This dataset included individuals aged 0 to 19 years, measured in months, from five countries (Brazil, South Korea, Mexico, England, and the United States), enabling a multiethnic analysis [9,48].

Initially, individuals with body mass and height values that were below −2SD or above +2SD within their respective age groups and sexes were excluded [9,48]. Body mass and height values were then transformed to a natural logarithmic scale to achieve maximum homoscedasticity in the distributions [9,48]. After-transformation, the data were fitted into a log–log linear regression model defined as log (body mass) = α x β (height) [9,48]. Subsequently, the p_t_ coefficients were calculated from the combined dataset of all five countries using a spline modeling technique (5 knots not determined a priori) [9,48].

Additionally, an analysis of variation among ethnic groups was conducted using mixed-effects models to investigate the intraclass coefficient of variation of individual height, body mass (calculated by the residual of linear regression of weight on height to estimate body mass not predicted by bone mass), BMI, and ABMI [48]. Overall, the mixed-effects analyses demonstrate that the greatest variability was observed in height measurements across different epidemiological contexts, while the ABMI showed a practically null variation among the studied ethnicities [48]. This suggested that ABMI was independent of height and showed no effect of contextual ethnic variability, highlighting its potential as an international index with p_t_ coefficients specified only for age and sex [48].

These p_t_ coefficients were applied for boys from 5 to 18 years old and for girls from 5 to 16 years old. For boys older than 18 and girls older than 16, the exponent of 2 was applied. The decision of applying the exponent of 2 for girls earlier than boys was made since studies point out that puberty occurs first in girls, which stabilizes their growth earlier, thus, there was no need to adjust their p_t_ coefficient after that [16,19,49].

The MULT ABMI-for-age growth charts were modeled from five to 22 years old and specified by sex (defined at birth by the presence or absence of a Y chromosome). This age group was chosen to ensure that the sample’s final height would be achieved since boys can still gain some height after the age of 18 [16,50,51]. MULT ABMI growth charts were constructed through the Generalized Additive Models for Location Scale and Shape (GAMLSS) approach, and the LMS method [52,53]. Moreover, the model class defined by the Box-Cox Cole Green distribution with penalized splines as the smoothing function was chosen for the distribution parameters L (skewness), M (median), and S (coefficient of variation) [53]. Considering that each penalized spline has a set of degrees of freedom (df) and can produce a variety of results based on the df range selection, the df of the L, M, and S parameters were selected using the Bayesian Information Criterion (BIC) to prevent overfitting [54,55]. For both sexes, the following df ranges were tested: L = 0-1, M = 0-4, and S = 0-3; then the one that presented the lowest BIC values was chosen (L=1; M=4; S=3). Furthermore, the Rigby and Stasinopoulos algorithm was used to fit the final model’s L, M, and S parameters [53,56].

The underweight, overweight, and obesity percentile cutoffs of the MULT ABMI growth charts were derived at ages 17, 18, 19, and 20 for girls and at ages 18, 19, and 20 for boys, using the values of 17 kg/m^2^, 25 kg/m^2^, and 30 kg/m^2^, respectively. The cutoff at 17 years old was estimated only for girls, since they stabilize growth and body mass before boys, suggesting that the cutoff can be established earlier for them [16,49,50]. The BMI values of 25 kg/m^2^ and 30 kg/m^2^ were selected because they were recommended by WHO as cutoff values to establish overweight and obesity in adults [3]. In addition, they were also used as cut-off values in DUTCH, WHO, IOTF, and MULT BMI references [22,24–26]. Similarly, the value of 17 kg/m^2^ was chosen to define underweight in our growth charts, since other BMI references, such as MULT, WHO, IOTF, and DUTCH, also use this value as a cut-off point for underweight [22,24–26].

The comparison between the CDC, WHO, IOTF, DUTCH, and MULT BMI and MULT ABMI growth charts was performed describing their trajectory based on the 50^th^ percentile (M curve), and the differences between BMI Z-scores from the other charts and ABMI Z-score were calculated, assuming ABMI as reference. Furthermore, the S and L curves, and the percentile curves for obesity established using the cutoff points at 18, 19, and 20 years old of these growth references have been described.

Concerning the L, M, and S curve model fit, the ABMI median, coefficient of variation, Box-Cox transformation parameter (L curve gross values), and worm plots developed by Van Buuren and Fredriks were applied [53,57]. All statistical analyses were performed in R version 4.2.1 for Windows and the RefCurve Software was used for estimating the MULT ABMI-for-age reference values [55,58].

## Results

After the exclusion of BMI value outliers and subjects who did not meet our eligibility criteria, 65 644 ABMI values (51.31% from boys) of 17 447 subjects (boys = 51.01%) were used in the MULT ABMI reference construction, as shown in Fig 1. The largest ethnic group of our sample was White (48.69%), followed by Asian Indian (14.10%), Black (13.39%), East and Southeast Asians (12.57%), and Native Peruvian (11.25%). The ABMI observations per sex and age groups are presented in Table 1.

**Fig 1.**
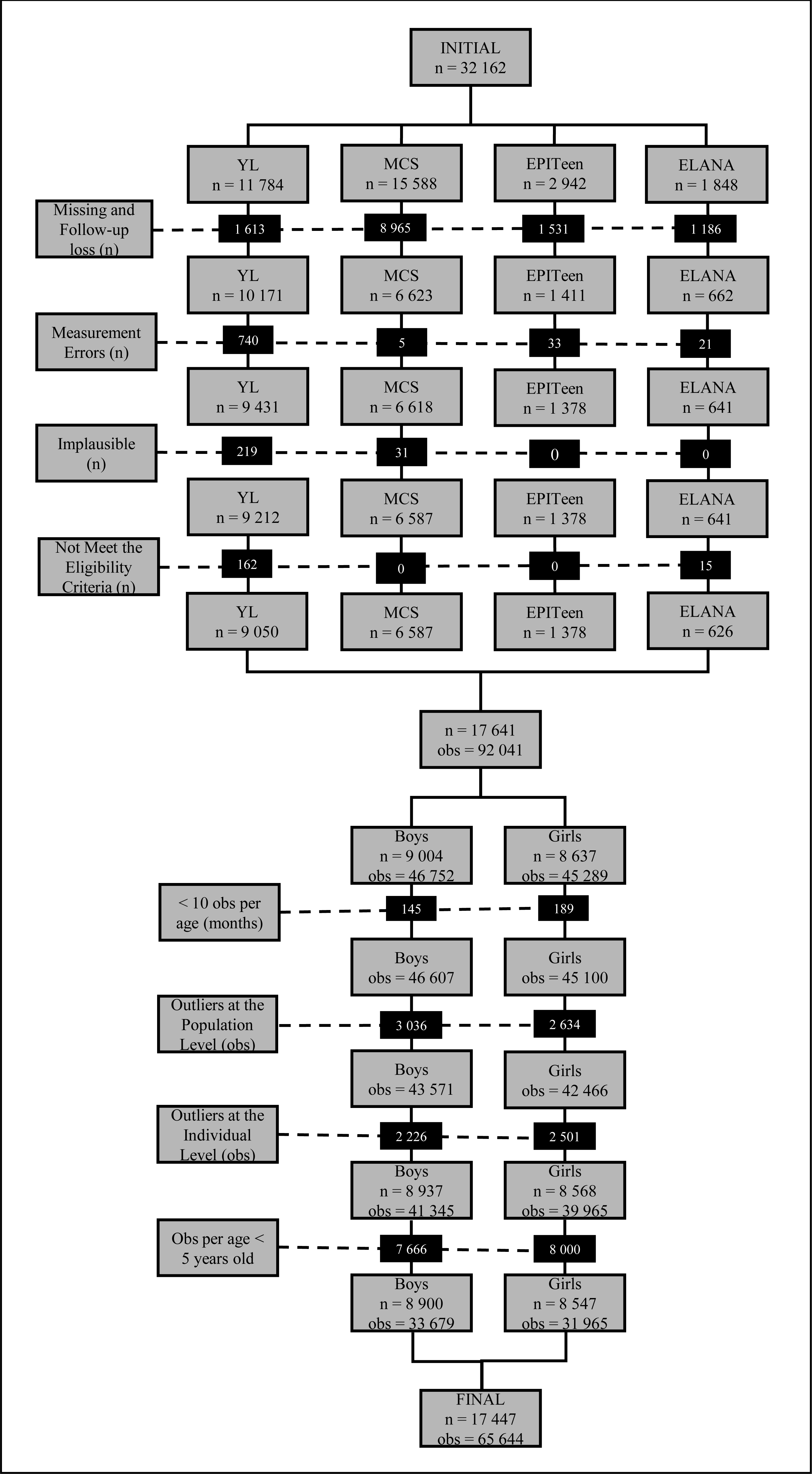
Flowchart of the subject and anthropometric data selection. n: number of participants obs: number of observations Measurement Errors: children and adolescents who had decreased their height over the years Implausible values: height-for-age z-score below –6 SD or above +6 SD or BMI-for-age z-score below –5 SD or above +5 SD. Outliers at the population level: weight-for-age z-score below −2SD or above +2SD (based on the WHO reference values for children under five years old and based on the weight of our sample for children from five years old and older. Outliers at the individual level: BMI-for-age z-score below −2SD or above +2SD (based on the BMI values of the linear mixed effects model).

**Table 1.**
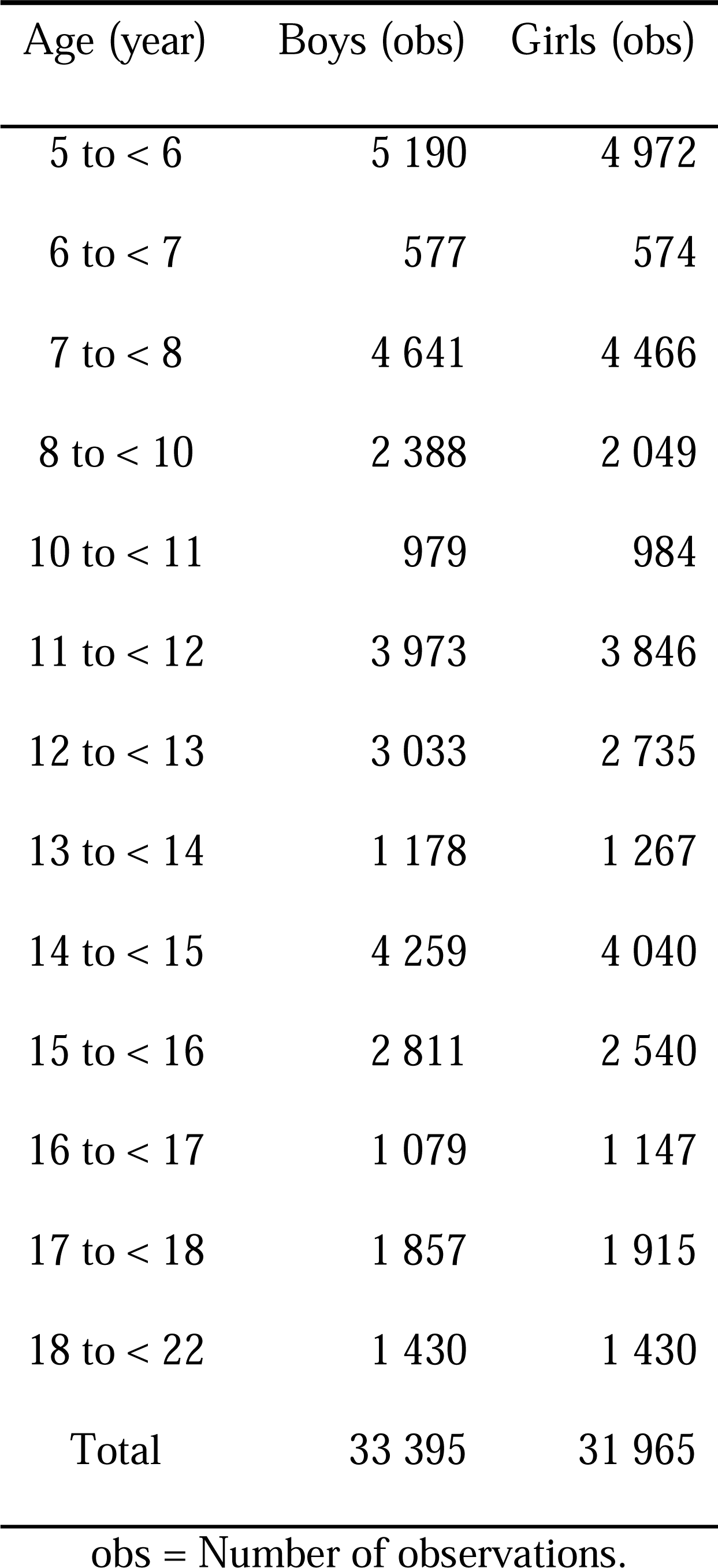
Number of observations per age group and sex included in the construction of the LMS parameters of the MULT ABMI reference.

The MULT ABMI LMS reference values and the ABMI p-coefficients (ABMI_p_t_) per age and sex proposed by Mazzeti et al. (2018) [9] are presented in Table 2. These coefficients were higher in girls than boys from 60 months to 84 months and between 108 to 132 months. On the other hand, at 132 months of age, the boys started to present higher p-coefficient values than girls, until 216 months (18 years old), when both had the isometric exponent (2), as shown in Fig 2.

**Fig 2.**
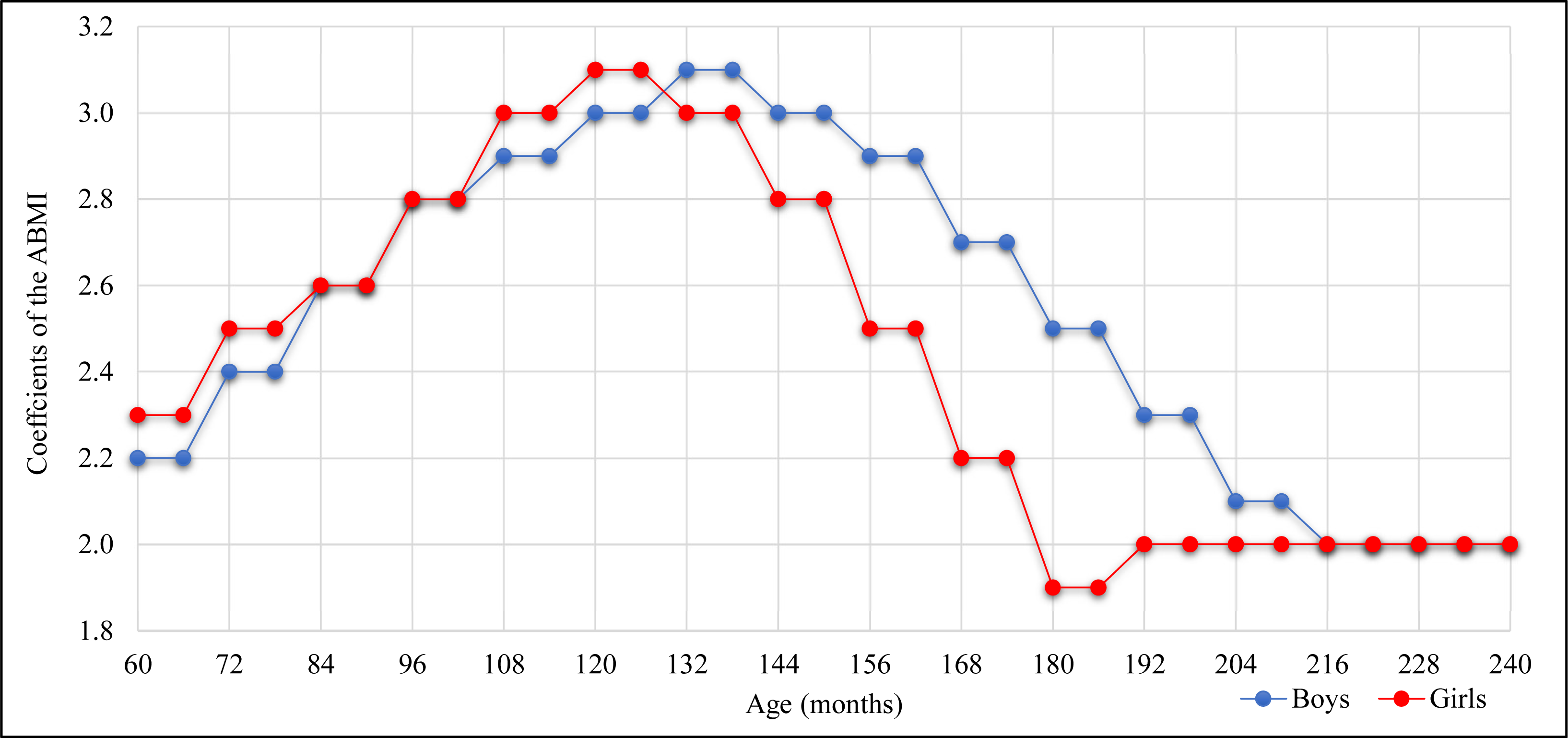
Allometric body mass index p-coefficient values for boys and girls. Coefficients of ABMI were previously estimated by Mazzeti et al. (2018) [9]. The comparison between the MULT ABMI and the other BMI references through the differences in body mass z-scores and their trajectories based on the M curve are shown in Fig 3. The MULT ABMI M values were lower than the all-BMI references from 64 to 234 months for boys and from 61 to 189 months for girls. The greatest differences between M curves were from 138 to 150 months for boys and from 114 to 132 for girls, which coincides with the puberty stage. The DUTCH and CDC BMI references presented the highest z-score differences to the MULT ABMI, while the MULT BMI and IOTF presented the closest values.

**Fig 3.**
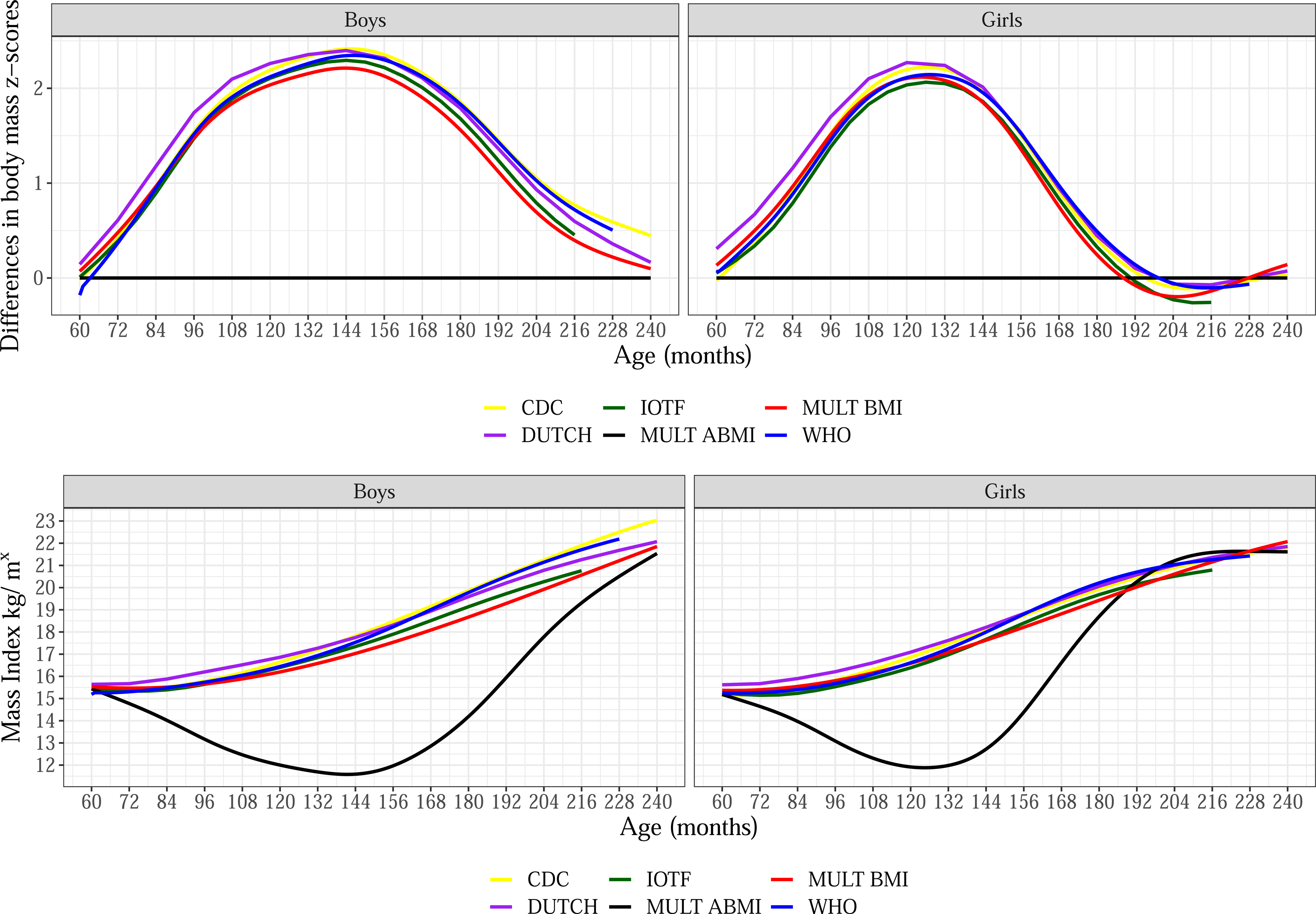
M curve and z-score differences of the five BMI references and the MULT ABMI reference in boys and girls. The S and L curves of the different growth references are shown in Fig 4. WHO S values were the lowest from 81 to 198 months, while MULT BMI S values were the greatest from 72 to 240 months. MULT ABMI presented S values similar to IOTF and DUTCH BMI references for boys from 60 to 114 months and then became higher, approaching the MULT BMI S values from 198 to 240 months. For girls, the MULT ABMI presented S values close to the IOTF, CDC, and DUTCH from 60 to 110 months, and then became higher, approaching the S values of MULT BMI until 240 months. Regarding the skewness, the L curve of MULT ABMI and BMI were more symmetric than the other BMI references, presenting the closest values to 0, while CDC presented the most asymmetric shape with values lower than −3.

**Fig 4.**
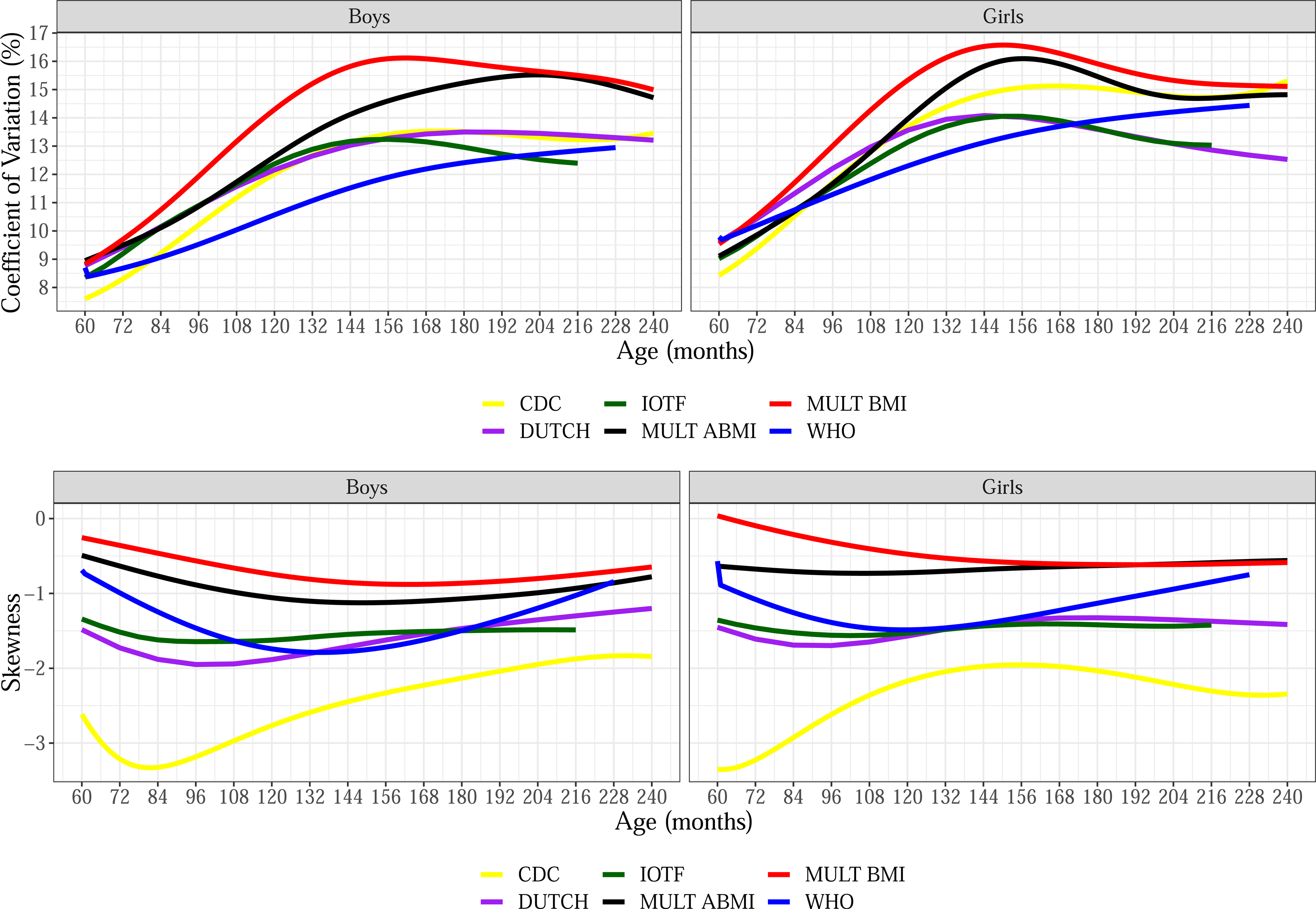
S and L curves of the five BMI references and the ABMI reference in boys and girls. In the comparison of the obesity percentile curves between the different references with cutoff points applied at 18 years old (216 months), the ones of MULT BMI and ABMI were below the IOTF and DUTCH BMI curves for both sexes, as shown in Fig 5. On the contrary, for the obesity percentile curves estimated applying the cutoff at 19 years old (228 months) the MULT BMI obesity percentile curve for the boys was above the WHO curve, while for girls it was very close until the age range of 156 to 216 months when the largest distance between these two obesity percentile curves is observed (Fig 5).

**Fig 5.**
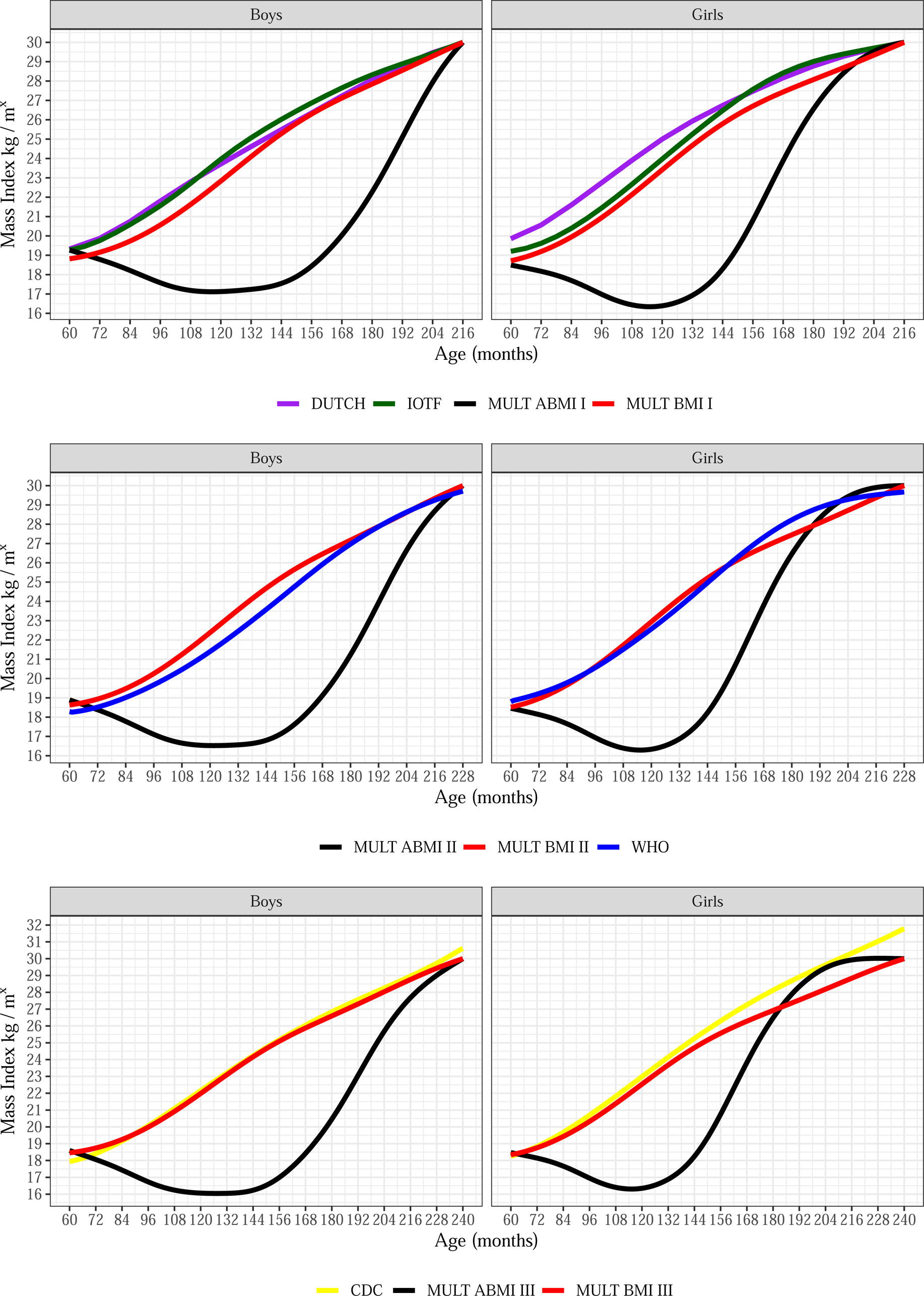
BMI percentile cutoffs for obesity at 18, 19, and 20 years old of the five BMI references and the ABMI reference in boys and girls. II - BMI value of 30kg/m^2^ applied as the cutoff for obesity at 18 years old III - BMI value of 30kg/m^2^ applied as the cutoff for obesity at 19 years old IV - BMI value of 30kg/m^2^ applied as the cutoff for obesity at 20 years old It is important to highlight that the MULT ABMI obesity percentile curve at 19 years old was below the others in the majority of the ages, surpassing the MULT BMI curve only for girls after 180 months of age and being above the WHO after 198 months. The MULT BMI and ABMI obesity percentile curves estimated by applying the cutoff points at 20 years old (240 months) were below the CDC curve for both sexes. The MULT ABMI growth charts for boys and girls are presented in Figs 6 and 7, respectively.

**Fig 6.**
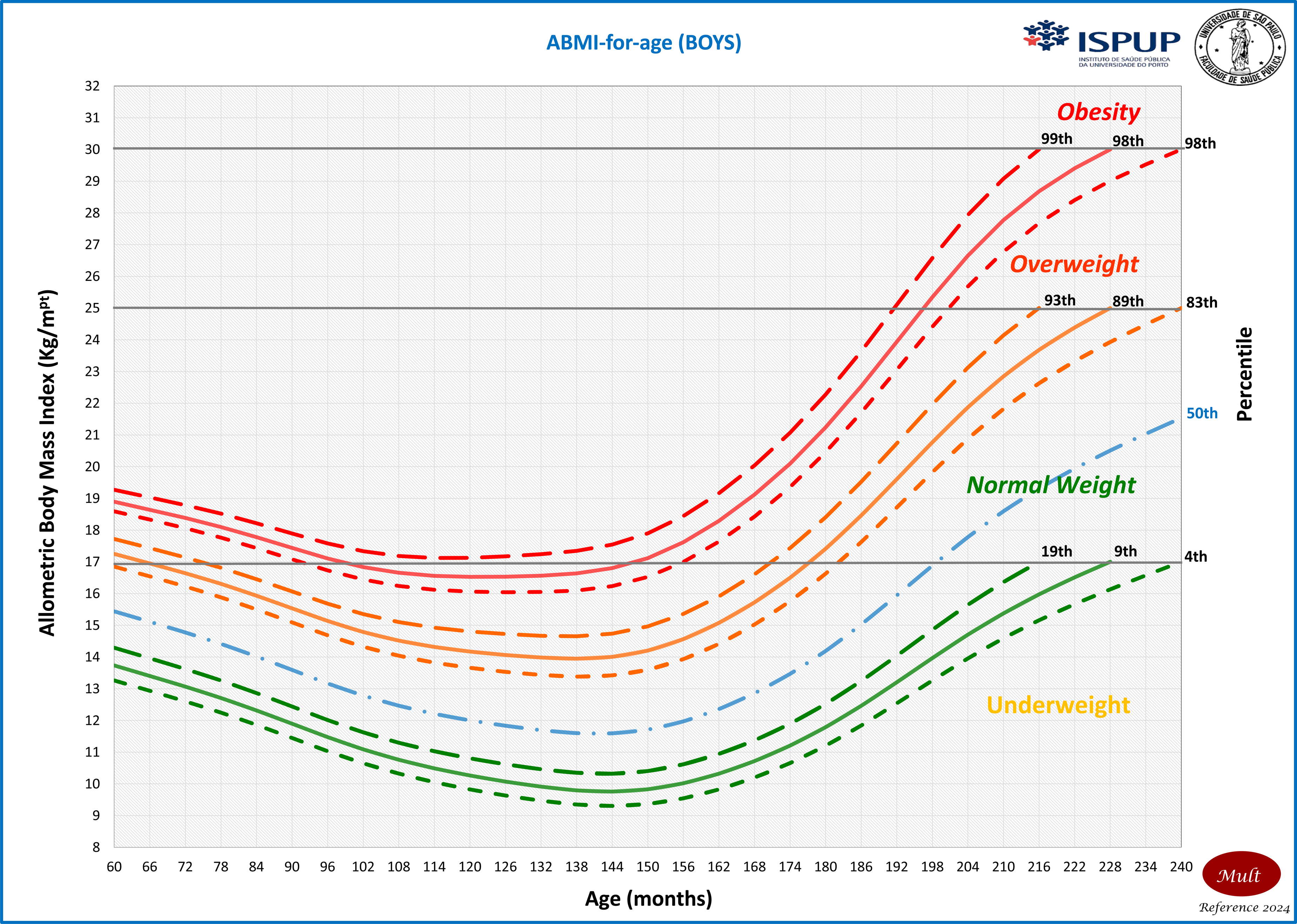
ABMI-for-age growth chart for boys aged 0 to 20 years old.

**Fig 7.**
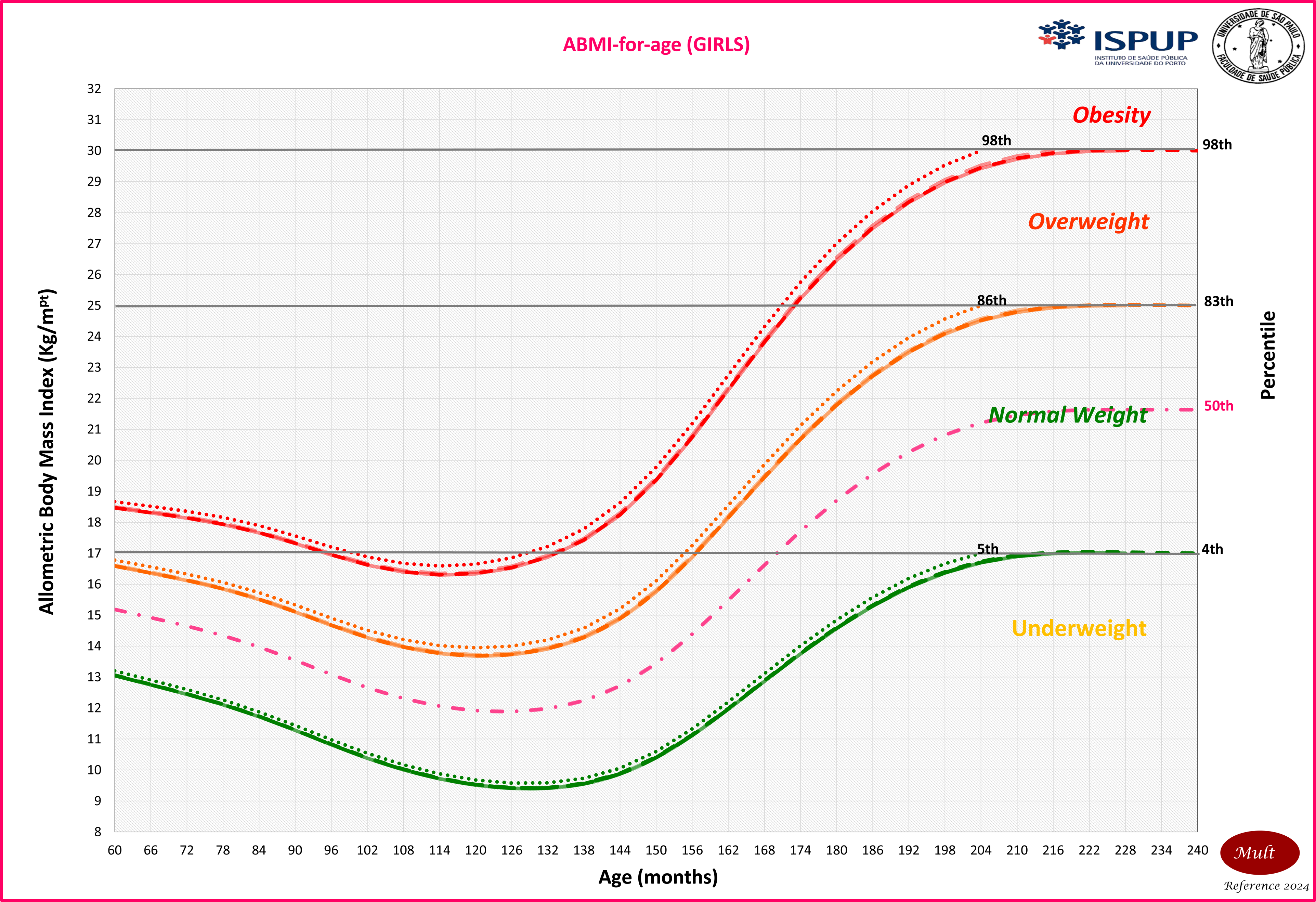
ABMI-for-age growth chart for girls aged 0 to 20 years old. Concerning the model fit of the MULT ABMI reference, the worm plots presented in S1 and S2 Figs showed that there were no deviations from the model’s assumptions, which indicates a good fit of the models.

**Table 2.**
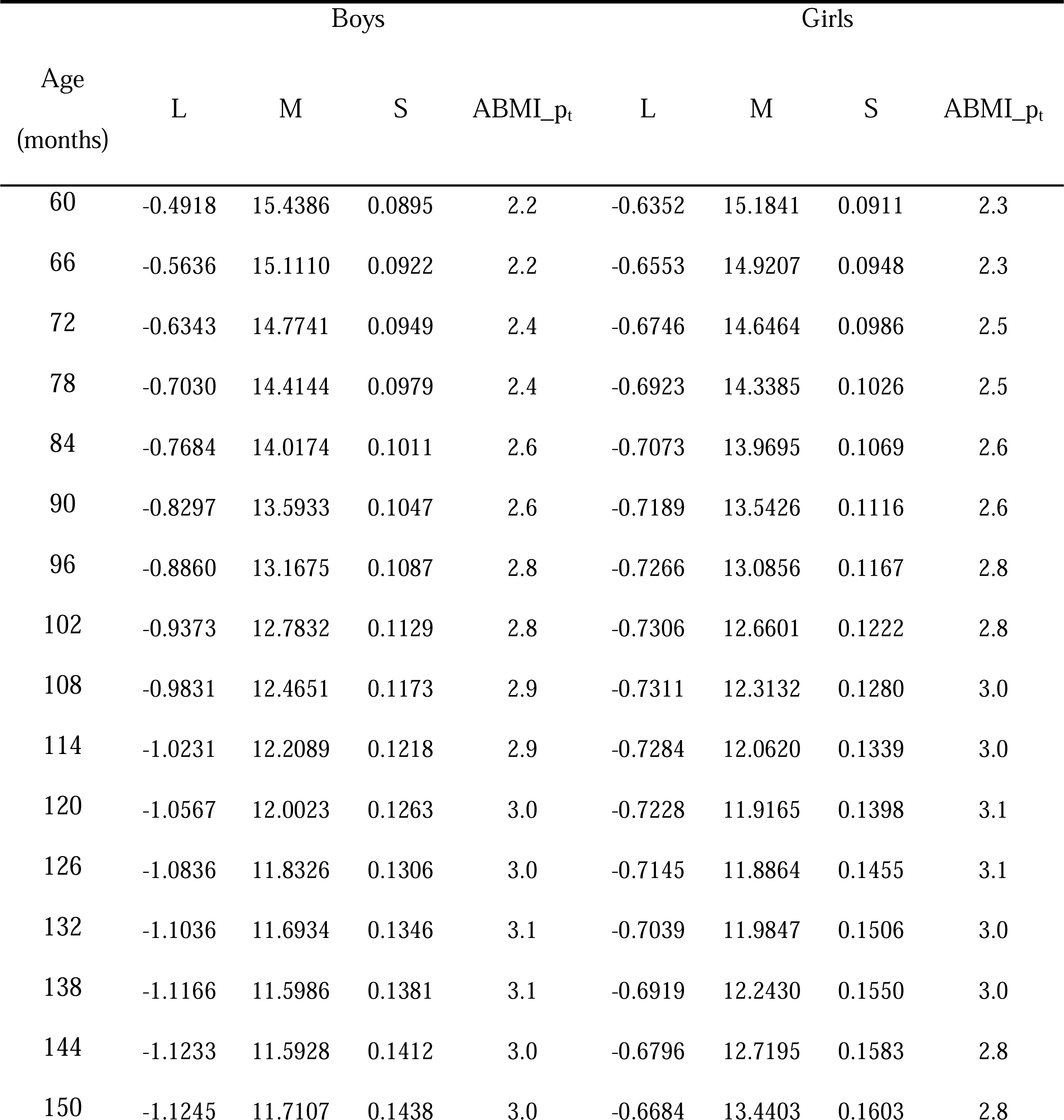
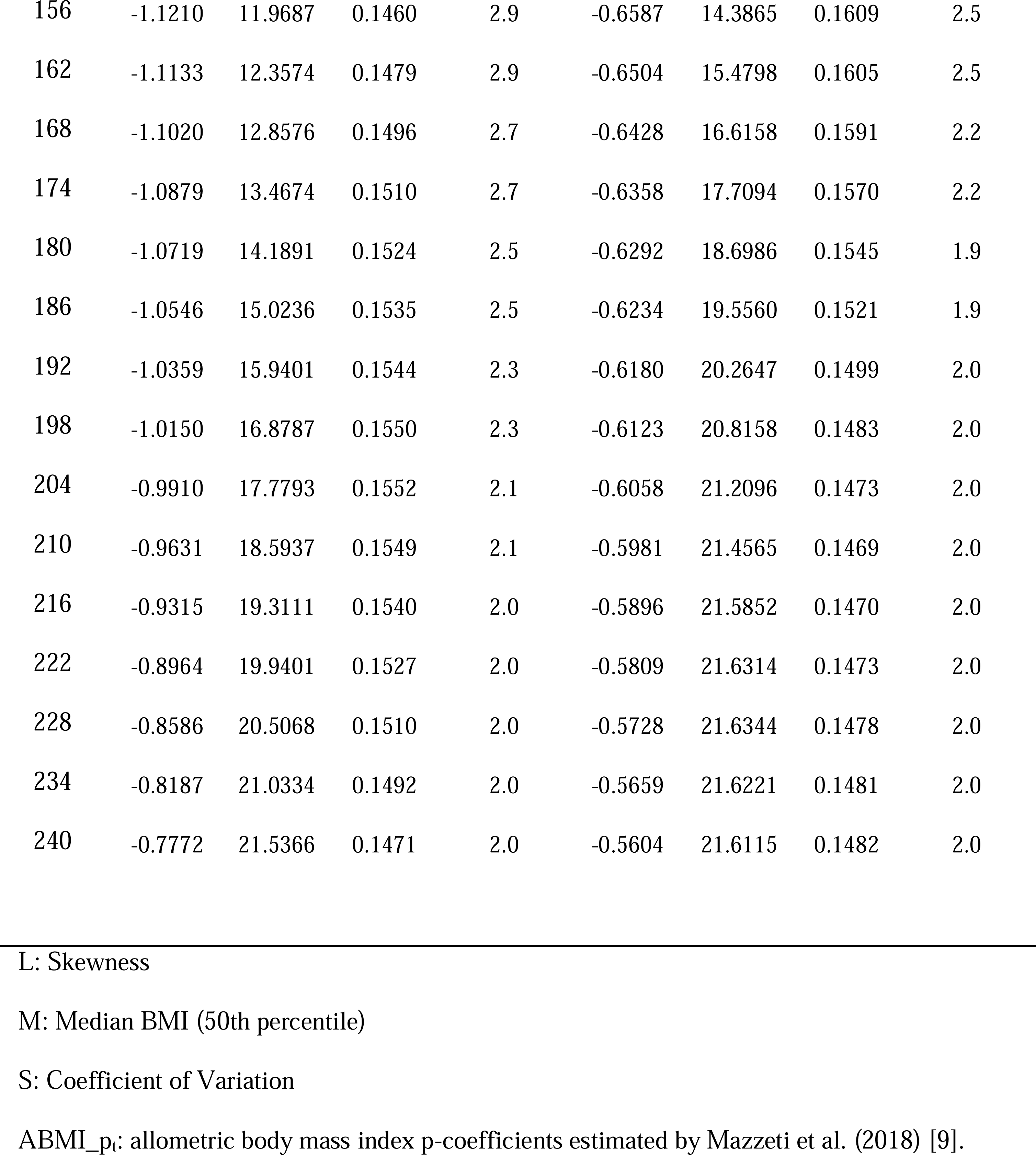
MULT ABMI LMS reference values and ABMI p-coefficients specified by age for boys and girls.

## Discussion

This study developed a MULT ABMI reference using allometric scaling to adjust the relation between weight and height. Several studies showed that indexes such as BMI (weight/height^2^) and ponderal (weight/height^3^) might not adjust well the relation between weight and height in some life cycle stages, and therefore this new reference would be a new approach to assess the nutritional status of children and adolescents [16–18,59].

BMI is widely used across populations, even though some researchers pointed out that the use of the same exponent to adjust the relation between weight and height for all age groups would not be adequate since there are changes in growth velocity and body composition during development [16,59]. From the first year of life until puberty, the weight gain increases slowly nearly as the square of the height, while during puberty there is an increase in the weight gain, stopping after age 25 years [4,16]. For this reason, Benn (1971) proposed an index with dynamic exponents that express the value for linearizing weight and height relation across age for each sex [8]. Accordingly, the exponent applied should provide the best correlation to weight and be uncorrelated with height [8]. This exponent should ensure the highest correlation between the index and adiposity combined with zero correlation between the index and height, being a value no higher than 3, and not necessarily an integer number [9].

During puberty, there is evidence that the exponent 2 applied in the BMI would not be enough to adjust the relation between weight and height [59]. Similar results were found by Cole (1986) that concluded that exponent 2 adjusts well the relation between weight and height for preschool children and adolescents after puberty, while during puberty this value should be increased to around 3 [16]. Additionally, Cole (1986) observed that, as a consequence of the pubertal process, girls experience their highest growth velocity 18 months earlier than boys [16]. This leads them to exhibit higher exponent values before boys, which adjusts the relation between weight and height [16]. A similar pattern was found by Mazzeti et al. (2018); in their study, the highest exponents were found in girls 12 months before the boys [9]. The explanation for that is that sexual maturation is the main factor for increasing this exponent, so as it occurs before in girls than boys, it is expected that they have the highest p-coefficients first [16,19,49]. Moreover, another study highlighted that height correlates to sexual maturation, therefore an adequate index should be independent of the height and correlated to adiposity as mentioned as an assumption of an ideal index [8,9,12,16].

These findings are important for understanding the premises of each index. For example, the BMI was not proposed as a way of predicting body fat, but rather as an index of the relation between weight and height [4,5]. The BMI’s association/correlation with obesity-related diseases has led to its wide use as a diagnostic tool for obesity screening [4,60]. For adolescents, the challenge is to have an index that adjusts the height in relation to the weight, taking into account the changes in growth velocity, body composition, fat, and weight gain and keeping the independence of the index from height, especially during puberty [61]. Unfortunately, BMI does not adjust it well during puberty, therefore the use of allometric scaling is suggested instead [9,16].

In the MULT ABMI reference rather than modeling the index, the fundamental relation between the weight and height during growth was modeled through the adjustments in the exponents, which were specified, by sex and age. During the 19^th^ century, Quetelet in his studies about body mass and growth indicated that in adolescents weight scales to height powered by at least 2.5 [5,62]. For this reason, correctly interpreting and use of body mass indexes have gained importance in the last years, especially to adjust weight and height to represent body mass or body fat independent of height [19,21,63–67].

Studies pointed out that the adjustment of this relation is important to obtain a BMI independent of height, an estimate of body fat regardless of the individual’s size, an indirect increase in the association between body mass and body fat, and the improvement of the precision and accuracy of nutritional diagnosis based on body mass [19,21,63–67]. Furthermore, the concept of shape is based on the premise that one dimension grows proportionally to the others and therefore its measurement is given by the ratio of two dimensions, which could be called volume [68]. According to Eveleth & Tanner (1991), this way of measuring shape can be an issue, as only two dimensions are considered and it can reduce the accuracy of the evaluation of the final index [68]. This is because, for example, dividing body mass by height does not take into account that the concept of shape is very close to the concept of volume, and would be necessary to place it in a quadratic or cubic scale [69,70].

In this way, considering the concept of body area and volume, the height measurement is intrinsically contained in the weight measurement, and to transform these two measurements into an index that assesses nutritional status, it is necessary to interpret that there are two vectors affecting growth and gain simultaneously [9]. These vectors are the chronological vector (linear growth and weight gain); and the biological vector (time, and translating growth velocity by different determinants) [13,14]. Therefore, in the BM analysis, the purpose is to use both proportions through an exponent, that could make the ratio independent of its denominator, regressing the measure of the numerator, which is the case of the BMI, calculated with the exponent equal to 2, for all individuals [9].

Regarding growth patterns, collecting data across diverse human populations, especially in small-scale populations, remains challenging [71]. In addition to that, there is an ongoing debate about the extent to which environmental factors influence these growth patterns [71]. For instance, studies conducted by Hruschka et al. (2019) and Wells (2017) suggest that some differences in height and weight cannot be explained mainly by environmental factors [72,73]. In the Hruschka et al. (2019) study these differences likely stem from a combination of unmeasured environmental and genetic factors [72]. Failing to consider these factors can lead to misinterpretations of phenotypic variations as indicators of socioeconomic and environmental disparities [72]. Wells (2017) pointed out that while adult height has a significant heritable component, the specific pathway through which it is attained also has substantial implications for metabolic phenotype [73].

Moreover, a study conducted with the Shuar, an indigenous Amazon population, revealed that their physical growth exhibited lifelong small stature and growth patterns that diverge significantly from the international standards/references of WHO (2006/2007) and CDC (2000), particularly after early childhood and around puberty [23,24,45,71]. This distinct growth pattern is due to the phenotypic plasticity and genetic selection in response to local environmental and life history factors, including a high burden of infectious diseases and high rates of mortality, which likely explain many aspects of Shuar growth [71]. Indigenous populations represent a significant portion of human genetic and cultural diversity but are often underrepresented in studies on childhood growth [74–76].

On the other hand, several studies have identified nutritional inadequacy and infectious diseases as the primary factors contributing to slow childhood growth and delayed maturation [44,77–79]. In this way, while some researchers have suggested that relying on a single international reference may not be appropriate for assessing growth worldwide, especially due to the varied growth patterns among non-Western and indigenous groups, it has been demonstrated that the lack of ethnicity diversity in these growth references may be the underlying issue [7,80–89].

Therefore, an advantage of the MULT ABMI reference is the inclusion of populations that have been underrepresented in the development of international growth references [26,51]. These populations include Africans (Ethiopians), South American Natives, various Asian groups (Indians and Vietnamese), and samples from multiethnic countries like Brazil, which exhibit a wide range of ethnicities [26,51]. Corroborating to it, some authors point out a need for combining data from different populations when estimating growth references to be used worldwide [25,45,80,90]. Another benefit was the use of longitudinal data, which considered the differences across the ethnic groups and allowed us to assess their growth trajectories over the years, especially during puberty [91].

Regarding the S curves, when compared to MULT growth references, WHO, CDC and IOTF presented lower coefficients of variation, especially during adolescence, a period when the growth spurt and the development of the genitals and secondary sex characteristics occur [49]. One possible explanation for that is the ethnic diversity lack in the IOTF, WHO, and CDC growth references [7,23,24,80]. WHO and CDC BMI references used data only from the United States, while in the IOTF BMI reference, African countries were absent in their sample population, as highlighted as a limitation of the IOTF growth charts [7,23,24,80]. As an example of that, a study conducted with school-age children pointed out that IOTF growth charts presented obesity misclassification for Ghanaian children in comparison with the deuterium dilution method [92].

Furthermore, another advantage of the MULT ABMI reference was to cover all the first years of early adulthood in its modeling, which was not done in the IOTF growth reference [80]. This is an important factor, since there is some evidence that boys grow even after 18 years old, and cutoffs established before 19 or 20 years can generate greater sex differences in the cutoffs [50,51,80,93]. Moreover, some authors suggest that the adult BMI cutoff points proposed by the WHO (2000) should be used only after 19 years old [3,24,94].

Concerning the underweight, overweight, and obesity cutoffs, the MULT ABMI reference presented options estimated from the BMI cutoffs at 17, 18, 19, and 20 years old. These ages are commonly applied as the upper limit for the international BMI growth charts [22–26,80,90]. Similarly, the BMI value of 30 kg/m^2^ is commonly applied to determine the obesity percentile cutoff in international BMI references [22,24–26,80,90]. This obesity cutoff point is well known as a risk factor for developing some types of cancers, non-communicable diseases such as diabetes, cardiovascular disease, and some respiratory, chronic musculoskeletal, infertility, and skin problems [3]. Another advantage of the MULT ABMI reference is to provide curves based on different cutoff options, because it generates the opportunity for each country to test the one that best suits its population according to its national growth patterns.

Regarding the strengths of this study, it is the first one to propose an international growth reference based on the ABMI, instead of using the BMI, which has limitations for the weight to height relation in some age groups. Additionally, we used standardized longitudinal surveys with a large sample and higher ethnicity diversity, as well as advanced techniques to predict the MULT ABMI reference values and to avoid outlier effects. Moreover, rather than self-reported data, the anthropometric data were gathered by trained experts, which is supposed to reduce the odds of measurement errors and social desirability bias [44]. To ensure data quality and model reliability, a large sample size with almost equal distributions of boys and girls, as well as the exclusion of the outlier measures, were used. Cole’s LMS approach, GAMLSS, and worm plots were also utilized, which are well-known techniques used in the growth reference constructions and supported by other studies [22,26,95,96].

Nevertheless, there are certain limitations in our study, including the absence of a health status evaluation and the fact that some samples, like those from Brazil and Portugal, were not national samples. However, BMI value outliers in the population and individual levels were removed to ensure a sample that had not been compromised by excessive weight, even if there was no evaluation of health status. Concerning the use of city data, EPITeen is a population-based study from the city of Porto, which presented similar patterns to the national data [30,97]. On the other hand, ELANA is comprised of a convenience sample of subjects from Rio de Janeiro [29]. This city has over 15% of immigrants, mostly from different Brazilian states, presenting demographic characteristics close to the national data [98,99].

## Conclusion

For the first time, an international ABMI reference (MULT) based on longitudinal data of multi-ethnic populations from ten countries was constructed and presented. The MULT ABMI reference presented values close to IOTF and MULT BMI references, which are growth charts constructed with multiethnic populations. The puberty stage was the period when the MULT ABMI M curve presented the highest disparity compared to the M curve of the other BMI references. Despite that, there are similarities in the S and L curves of these growth references. These findings highlighted that the MULT ABMI reference keeps the essential similarities to the international BMI references, with the exception of the pubertal period when the exponent of the index seems to adjust the relation between weight and height better, supporting that the MULT ABMI reference could be a useful tool to assess the nutritional status of children and adolescents worldwide.

## Supporting information

S1 Fig. Worm plot of the ABMI reference for boys.

S2 Fig. Worm plot of the ABMI reference for girls.

## Data Availability

The data that support the findings of this study are available from the UK Data Service, ISPUP, UERJ, and UFRJ. Restrictions apply to the availability of these data, which were used under license for this study. MCS and YL data are available from the UK Data Service online platform at URL [https://ukdataservice.ac.uk] and EPITeen and ELANA data are available from the authors with the permission of ISPUP, and UFRJ, respectively.

## Acknowledgements

The authors are grateful to children and families who participate in the YL, MCS, ELANA, and EPITeen studies and to the UK Data Service, the UERJ, the UFRJ, and the ISPUP for making these datasets available to researchers. The authors would also like to thank the Department of Pediatric Cardiology of the University of Bonn for making the RefCurv software free to the scientific community.

## Supporting information

**S1 Fig. Worm plot of the ABMI reference for boys.**

**S2 Fig. Worm plot of the ABMI reference for girls.**

