## Supplementary figures and images for "MULT: An allometric body mass index (ABMI) reference to assess nutritional status of multi-ethnic children and adolescents"

### S1 Fig. Worm plot of the ABMI reference for boys.

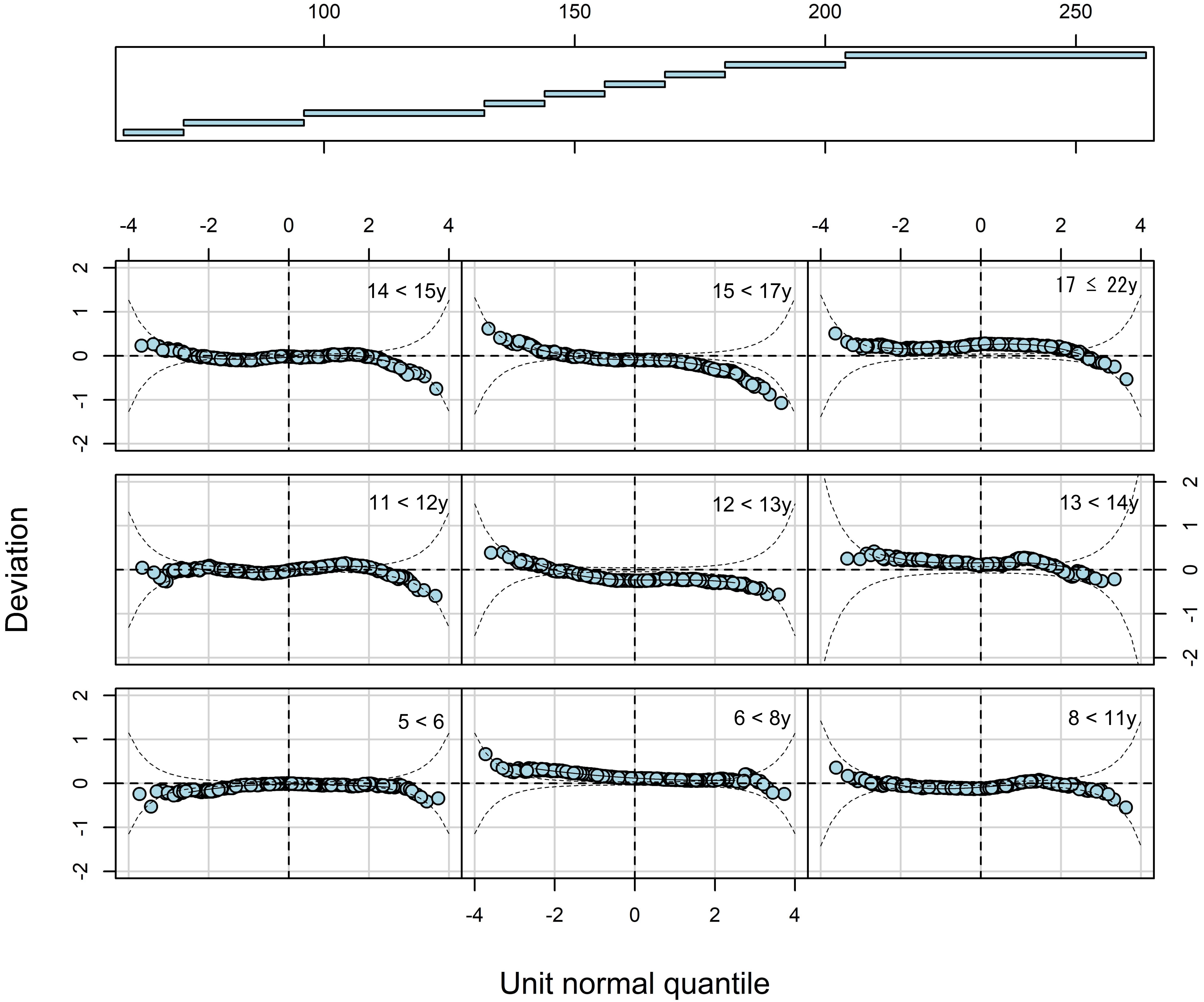

### S2 Fig. Worm plot of the ABMI reference for girls.

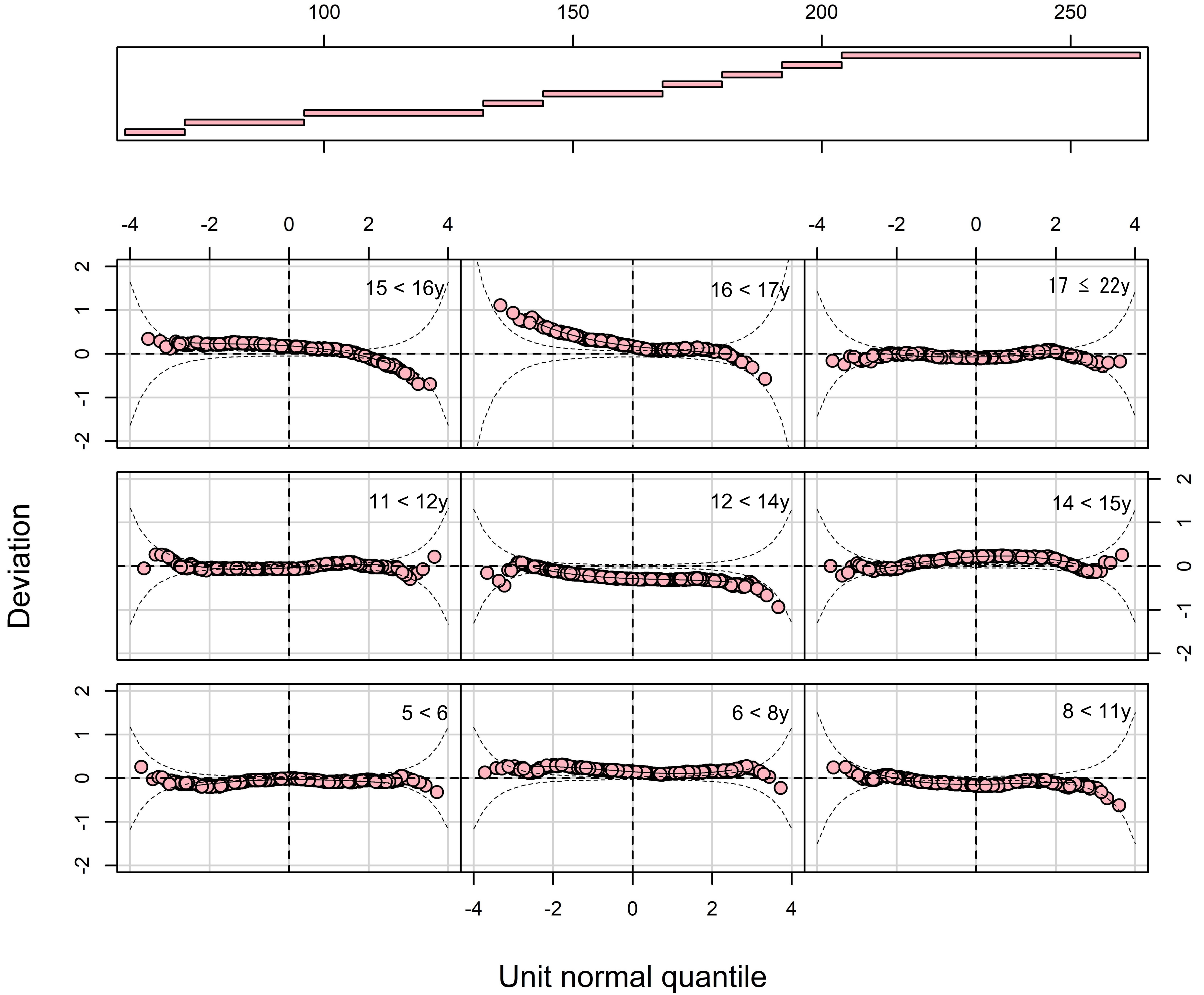
